# Personalizing Suicide Risk Assessment: Machine Learning Extraction of Cross-Modal Interactions Between Psychosocial and Demographic Factors in Veterans^1^

**DOI:** 10.64898/2026.06.16.26355796

**Authors:** Maxwell Levis, Brian Shiner, Monica Dimambro, Luke Rozema, Siamack Ayandeh, Alos Diallo, Yefan Zhou, Siting Li, Weiyi Wu, Jiang Gui, Joshua Levy

## Abstract

**Background:** Veterans face an elevated risk of suicide compared to the general population, motivating national efforts to develop predictive models that can guide proactive care. Current models used by the U.S. Department of Veterans Affairs (VA) rely primarily on structured electronic health record (EHR) data, though clinical notes contain rich contextual information that can be quantified using natural language processing (NLP) to derive psychosocial variables that may improve risk detection. Machine learning methods, particularly classification and regression trees (CART), can also uncover interactions between clinical and psychosocial variables, enabling identification of patient characteristics that modify suicide risk factors. However, integrating structured and unstructured data presents challenges because NLP features often greatly outnumber traditional clinical variables, potentially biasing interaction discovery. In prior work, we addressed this imbalance by introducing a weighted CART framework that balances structured variables with NLP-derived psychosocial features from semantic lexicons (SÉANCE). While effective, semantic approaches summarize language into predefined constructs and may overlook important lexical variation present in clinical narratives.

**Methods:** In this study, we extend that framework by replacing semantic features with a high-dimensional bag-of-words (BoW) representation of clinical notes and by evaluating models across cohorts defined by structured suicide risk stratification (low, medium, high) and varying temporal lookback windows. Using a cohort of 27,241 veterans, we analyzed clinical documentation collected up to 30, 90, or 270 days prior to death (or a matched index date for controls), enabling temporally flexible risk modeling. XGBoost models were trained to balance structured and unstructured features and identify cross-modal interactions between textual and clinical variables.

**Results:** When incorporated into generalized linear models, these interactions improved predictive performance, particularly among low- and medium-risk patients, and substantially reduced the performance gap between interpretable and more complex models. Notably, the BoW representation outperformed our prior semantic index–based approach.

**Discussion and Conclusions:** Together, these findings demonstrate the utility of interpretable NLP methods for uncovering clinically meaningful interactions between psychosocial and demographic factors in suicide risk and establish a strong benchmark for future deep learning approaches aimed at capturing richer contextual and temporal information from clinical narratives.

## 1. Introduction

Veterans face alarmingly high suicide rates, rates that are substantially higher than civilian populations ^1^. Addressing this urgent concern, the U.S. Department of Veterans Affairs (VA) has prioritized developing personalized suicide risk prediction tools. The VA’s model, Recovery Engagement and Coordination for Health – Veterans Enhanced Treatment (REACH-VET) is an algorithm which leverages structured electronic health record (EHR) data to identify and categorize individuals at elevated suicide risk ^2,3^. This system plays a vital role in identifying VA patients who are most at risk for suicide and connecting them with prevention services ^4^.

Recent studies have demonstrated that REACH-VET’s predictive accuracy can be enhanced by incorporating unstructured EHR data, such as clinical notes, to identify novel risk and treatment factors ^5,6^. Our earlier work used this methodology to evaluate risk typologies across different suicide risk-tiers ^7,8^, identifying Natural Language Processing (NLP)-derived variables that supplement traditional demographic and structured predictors. As part of these analyses, we used pre-established semantic indexes (Sentiment Analysis and Cognition Engine– SÉANCE) to convert note text into psychosocial variables related to risk and treatment, and evaluated strategies to integrate these features with structured clinical variables across REACH-VET-defined suicide risk tiers ^9,10^. Those studies emphasized the interpretability of interactions between SÉANCE variables and patient characteristics, using a novel weighting scheme to address imbalances in variable set sizes during classification and regression tree (CART) modeling. However, semantic indexes can introduce bias and may fail to capture the full lexical and contextual variability present in unstructured clinical text ^11–13^.

In the present study, we build on this foundation by adopting a more flexible and data-driven NLP approach using bag-of-words (BoW) representations, further enabling direct exploration of interactions between specific lexical features and patient characteristics. Rather than relying on pre-established semantic indexes, this method uses contextual language features derived from clinical notes to generate risk- and treatment-related variables that are both risk-tier-specific and time-sensitive. These variables are integrated with structured data using our previously introduced cross-modal weighting approach, yielding interpretable models that can uncover personalized risk signatures across subpopulations.

While deep learning methods hold promise for modeling nuanced language patterns, especially at scale, we view them as a future direction for this line of work. Given the clinical importance of interpretability and the relatively limited exploration of cross-modal interactions in this space, our focus here is on establishing principled methods using interpretable features. These approaches serve as an essential foundation before pursuing and comparing more complex architectures in subsequent studies.

## 2. Methods

### 2.1. Patient Selection, Risk Stratification, and Clinical Note Retrieval within Relevant Time Windows

Similar to our previous work, we constructed a retrospective matched case-control cohort using the U.S. Department of Veterans Affairs (VA) Corporate Data Warehouse (CDW), which contains nationwide electronic health records (EHR), in combination with mortality data from the VA–Department of Defense Mortality Data Repository ^14^. Veterans who died by suicide and had documented interaction with VA healthcare during 2017 and 2018 were identified, yielding 4,584 cases.

Each suicide case was matched to unique control patients who were alive at the time of the case’s death, yielding 22,657 controls (**Table 1**). Controls were selected to match on VA facility, calendar time, and REACH-VET risk percentile. No control patient was matched to more due to care site or baseline clinical risk and allows us to evaluate predictive performance of our models relative to REACH-VET.

**Table 1.** Patient Characteristics for parent 2017-2018 cohort with clinical notes extracted from one year time to death (used to derive 30, 90 and 270 day subcohorts); omitted variables relating to eligibility, disability, physical/mental health burden, select mental health risks, inpatient/ER, select prescriptions at index date

|  | Case<br>(N=4584) | Control<br>(N=22657) | SMD |
| --- | --- | --- | --- |
| **Demographics** |  |  |  |
| Female | 163 (3.6%) | 1565 (6.9%) | 0.151 |
| Married | 1931 (42.1%) | 10268 (45.3%) | 0.064 |
| Homeless Prior 2 Years | 60 (1.3%) | 318 (1.4%) | 0.008 |
| Veteran | 4542 (99.1%) | 22495 (99.3%) | 0.022 |
| Rural | 60 (1.3%) | 318 (1.4%) | 0.006 |
| Age: Mean (SD) | 61.0 (18.5) | 60.6 (16.5) | 0.024 |
| **Race** |  |  |  |
| Native American / Asian / Pacific Islander | 101 (2.3%) | 505 (2.2%) | 0.259 |
| Black | 240 (5.2%) | 2399 (10.6%) |  |
| Hispanic | 184 (4.0%) | 1355 (6.0%) |  |
| White | 3774 (82.3%) | 17692 (78.1%) |  |
| **Risk Tier** |  |  |  |
| High | 451 (10.2%) | 2205 (10.2%) | 0.948 |
| Medium | 2091 (47.4%) | 10306 (47.7%) |  |
| Low | 2042 (42.4%) | 10146 (42.1%) |  |
| **Deployment** |  |  |  |
| Vietnam | 1671 (36.5%) | 9167 (40.5%) | 0.084 |
| Afghanistan or Iraq | 1541 (33.6%) | 7931 (35.0%) | 0.037 |
| **Mental Health Risk Flag** |  |  |  |
| Anxiety | 1835 (40.0%) | 9187 (40.5%) | 0.011 |
| Bipolar | 725 (15.8%) | 2841 (12.5%) | 0.094 |
| Depression | 2649 (57.8%) | 12744 (56.2%) | 0.031 |
| PTSD | 1451 (31.7%) | 7161 (31.6%) | 0.001 |
| Sleeping | 1865 (40.7%) | 10023 (44.2%) | 0.073 |
| Substance | 1779 (38.8%) | 7374 (32.5%) | 0.131 |
| Trauma | 1985 (43.3%) | 9986 (44.1%) | 0.016 |
| **Prescriptions Prior 12 Months** |  |  |  |
| Opioid Rx | 1073 (23.4%) | 5202 (23.0%) | 0.011 |
| Mood Stabilizer Rx | 1263 (27.6%) | 5835 (25.8%) | 0.041 |
| Antipsychotic Rx | 748 (16.3%) | 2838 (12.5%) | 0.108 |
| Antidepressant Rx | 2000 (43.6%) | 9887 (43.6%) | 0.000 |

To account for heterogeneity in suicide risk presentation, patients were stratified into three REACH-VET risk tiers (low, medium/moderate, and high) prior to model development. These were defined in earlier work as REACH-VET scores of 100-25 (low), 24-1 (medium), and 1 (high), respectively (**Figure 1A**) ^8^. Within each risk tier, we retrieved unstructured clinical notes from the CDW written in the <30, <90, and <270 days leading up to each case’s death (or matched index date for controls) (**Figure 1B**). Because inclusion in each time window required at least one eligible note within that interval, some patients were excluded from shorter windows (e.g., <30 days) if all their notes fell entirely outside the specified range. In contrast to our prior work, which excluded only notes from the final two days ^15^, we implemented a more conservative exclusion to reduce endogeneity by removing all notes recorded within six days of the index date. This approach ensured that modeling was based on contemporaneous clinical documentation not likely influenced by imminent death.

**Figure 1.**
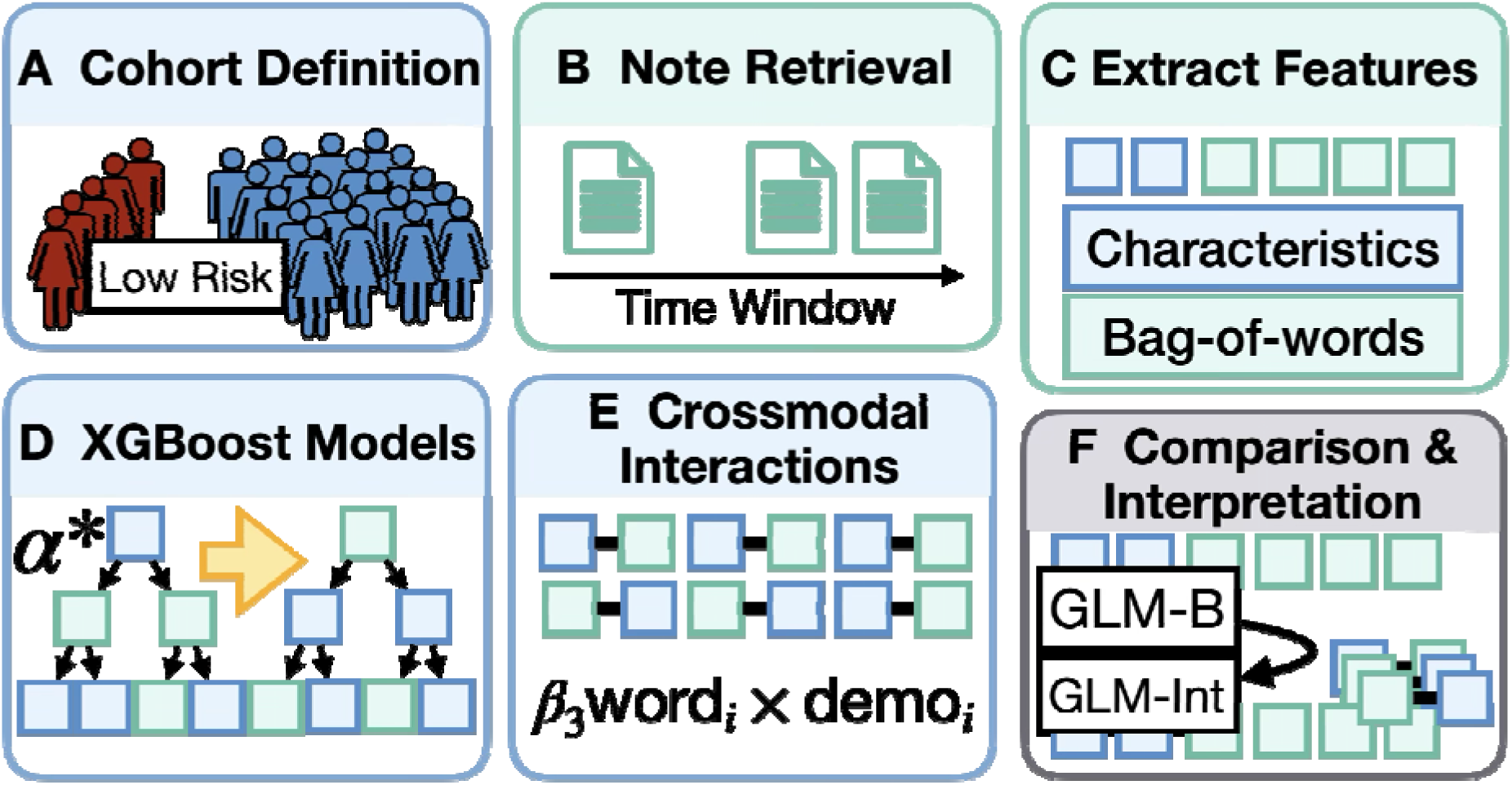
Method Overview. **A)** Patient cohort defined through REACH-VET matching of cases and controls, further stratifying patients within EHR/REACH-VET defined risk tiers (low, medium/moderate, high). The low-risk tier is depicted as an example subgroup; the workflow was applied separately within all risk tiers. **B)** Clinical notes retrieved within defined time intervals from death or index event (<30, <90, <270 days). **C)** Clinical/demographic and text bag-of-words count NLP features extracted; number of words far outnumbers number of clinical features. **D)** XGBoost models are fit which fairly balance sampling of clinical versus NLP features. **E)** SHAP used to extract cross-modal interactions between clinical and NLP features; interactions validated through negative binomial regression. **F)** Identified interactions are added as additional features in logistic regression (generalized linear model; GLM) approach (GLM→GLM-Int; both fit using elastic net followed by L2 penalization) to assess the overall impact and performance improvements attributed to the cross-modal interactions; select interactions are interpreted using estimated marginal means

To further avoid overrepresentation of high-utilization patients, individuals with more than six times the average number of notes within an interval were excluded. This resulted in a dataset of 378,697 notes from 1,934 cases and 9,702 controls at low risk, 859,075 notes from 2,055 cases and 10,105 controls at moderate risk, and 566,710 notes from 449 cases and 2,198 controls at high risk (**Figure 1B**). **Table 2** summarizes the number of suicide cases, matched controls, notes, and vocabulary size for each combination of risk tier and time window. Modeling and interpretation were conducted at the note level, while performance evaluation was performed at the patient level based on aggregated predictions.

**Table 2.** Data Overview. Summary of Patients, Case-Control, Notes, Vocabulary Size by Risk Tier and Days-to-Death. Size of vocabulary denotes the number of unique unigrams and bigrams in the respective corpora.

| Risk Tier | Days to Death | Patient Count | Cases | Controls | Note Count | Size of Vocabulary |
| --- | --- | --- | --- | --- | --- | --- |
| **Low** | <270 | 11636 | 1934 | 9702 | 378697 | 3186 |
|  | <90 | 8960 | 1473 | 7487 | 126776 | 3198 |
|  | <30 | 5677 | 953 | 4724 | 38493 | 3308 |
| **Moderate** | <270 | 12160 | 2055 | 10105 | 859075 | 3154 |
|  | <90 | 10762 | 1794 | 8968 | 306292 | 3145 |
|  | <30 | 8099 | 1364 | 6735 | 90092 | 3084 |
| **High** | <270 | 2647 | 449 | 2198 | 566710 | 3279 |
|  | <90 | 2550 | 431 | 2119 | 247874 | 3354 |
|  | <30 | 2263 | 376 | 1887 | 90874 | 3309 |

### 2.2. Data Extraction and Preparation

#### 2.2.1. Clinical note data collection and preprocessing

For each clinical note, we applied standard text preprocessing steps: lowercasing, punctuation removal, and the elimination of common stop words (e.g., “ the”, “and”, “was”). Text was then tokenized into both unigrams and bigrams. Using Scikit-learn ^16^, we computed both term frequency–inverse document frequency (TF-IDF) and raw count-based representations. To reduce vocabulary size and minimize sparsity, we retained only terms that appeared in at least 1% and at most 90% of documents within each stratum (risk tier × time window). This heuristic was selected to balance the inclusion of salient terms while mitigating excessive zero inflation in downstream modeling. Although both TF-IDF and count vectors were computed, we ultimately used raw count representations for all modeling, as they allow for more interpretable associations—particularly with negative binomial models used for interaction validation (see Section 2.4). The resulting vocabulary sizes varied across risk tier and time window combinations, ranging from 3,084-3,354 unigrams/bigrams (**Table 2**, **Figure 1C**).

#### 2.2.2. Extraction of Structured Patient Characteristics

Structured variables were extracted from the VA CDW to capture a broad profile of patient demographics, health status, and service utilization (**Table 1**, **Figure 1C**). These included core demographic features (e.g., age, sex, race, marital status) and veteran-specific demographics such as military service branch, as well as indicators of social vulnerability, such as homelessness history. We also incorporated measures of healthcare engagement, including the number and types of visits to emergency departments, primary care, and mental health services. Medication history was derived from pharmacy records, highlighting prescriptions for opioids, antipsychotics, and other relevant drug classes. Diagnostic data was identified by ICD code and spanned a range of mental health conditions such as PTSD, depression, anxiety, substance use disorders, and bipolar disorder (**Table 1**). Multivariate Imputation by Chained Equations (MICE) was used to impute any missing variables for an insignificant subset of observations from the entire study cohort (2.4% of patients were missing five or more variables) ^17^. This resulted in a total of 61 structured variables, consistent with our prior work ^10^, which served as the input feature set for the demographics-only and combined models.

#### 2.2.3. Dataset Partitioning Strategy

To support model development and evaluation, we partitioned patients into training (64%), validation (16%), and test (20%) subsets for each combination of risk tier and time window. This split was performed using the StratifiedGroupKFold function from the Scikit-learn library (Python v3.9) ^16^, which enabled stratification while preserving group membership at the patient level. All clinical notes and structured variables for a given individual were confined to a single partition, thereby preventing any overlap across sets and reducing the risk of data leakage. This patient-level grouping ensures that performance estimates on the held-out test set are not artificially inflated by exposure to related data during training.

### 2.3. Modeling Framework and Cross-Model Feature Integration

#### 2.3.1. Prioritizing Crossmodal Interactions During XGBoost Model Training

We trained gradient-boosted tree models using XGBoost to classify suicide decedents across each combination of suicide risk tier and temporal interval (<30, <90, <270 days) ^18^. Three model variants were considered for each stratum (**Figure 1D**):

1. **XGBoost (Demo):** Structured variables only (demographics and clinical features),
2. **XGBoost (Count):** Unstructured language variables only (bag-of-words/BoW features), and
3. **XGBoost (**α**\*):** A weighted combination of both modalities, using a tunable mixing parameter α ∈ [0, 1] to control the relative sampling probability of features from each set.

The α-weighting strategy was introduced in our previous work ^10^, where we showed that conventional tree-based models may under-sample smaller feature sets (e.g., patient characteristics) when competing with high-dimensional text data. Controlling variable sampling probabilities (α) allows us to explore performance tradeoffs and interaction discovery under different integration regimes. Briefly, the sampling probabilities 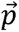 for each *jth* patient characteristic is 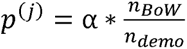 and for each unique *jth* unigram/bigram is *p*^(*j*)^ =1 - α, then normalizing all feature sampling probabilities such that 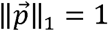. The feature set sizes, *n_BoW_* and *n_demo_*, reflect the number of unstructured and structured predictors, respectively, as described in *sections 2.1-2.2*.

For the present study, we selected the optimal α for each stratum based on validation AUROC and used the resulting models for downstream interaction analysis and performance evaluation. All XGBoost models were fit using a randomized grid search over hyperparameters, with early stopping based on validation set performance (**Table 3)** ^19–21^.

**Table 3.** Hyperparameters– for 50-iteration randomized grid search. A consistent random seed was applied to ensure the same hyperparameter set was evaluated across risk tiers and time windows. **Note:** The α cross-modal weighting parameter used in XGBoost is optimized separately and is not included here.

| Model | Hyperparameter | Values |
| --- | --- | --- |
| **GLM** | 1/L2-Penalty | [0.0001, 0.001, 0.01, 0.1, 1, 10] |
|  | L1 Penalty | 0.2 (fixed after coarse search) |
| **XGBoost** | Sampled columns by node | [0.25, 0.50, 0.75, 1.00] |
|  | Sampled columns by level | [0.25, 0.50, 0.75, 1.00] |
|  | Sampled columns by tree | [0.50, 0.75, 1.00] |
|  | Sample subsampling | [0.2, 0.4, 0.6] |
|  | Minimum child weight | [1, 3, 5, 7] |
|  | Maximum tree depth | [3, 4, 5, 6] |
|  | Gamma | [0, 1, 5, 10] |
|  | Regularization (L1) | [0.0, 0.1, 1.0, 10.00] |
|  | Regularization (L2) | [0.0, 0.1, 1.0, 10.00] |
|  | Number of Trees | [25, 50, 100] |

#### 2.3.2. Penalized Generalized Linear Models (GLMs)

To evaluate the contribution of cross-modal interactions within an interpretable framework, we constructed two regularized logistic regression models, fit with stochastic gradient descent (SGD):

1. **Baseline GLM (GLM-B):** A penalized logistic regression model using only main effects from structured (demographic) and unstructured (word count) features.
2. **Interaction-Augmented GLM (GLM-Int):** Using the *interactiontransformer* package, SHapley Additive exPlanations (SHAP) interaction scores were used to identify suicide-related cross-modal interaction terms between structured and unstructured features (**Figure** 1E, see Section 2.4) ^22,23^. These interaction terms were added to the baseline GLM-B model to create the interaction-enhanced model (GLM-Int). To ensure proper model specification and interpretability, any feature appearing in an interaction term but not already included among the GLM-B main effects was also added to the model as a main effect.

Both models were trained using the same two-stage modeling pipeline. In the first stage, Elastic Net regularization (combining L1 and L2 penalties) was applied for feature selection across the full predictor set ^24^. The top 150 predictors, determined through grid-search hyperparameter tuning of the L1 and L2 penalties (assessed using validation set AUROC), were retained. In the second stage, focused on predictive performance, selected features (with or without interactions) were passed to a ridge logistic regression classifier (L2-penalized), with a separate hyperparameter sweep to optimize validation-set performance. This framework allowed us to evaluate the overall contribution of interaction terms as a group, while reserving more parsimonious interpretation of individual effects for subsequent analyses. This process was repeated for each risk tier and time window configuration. All continuous structured variables were standardized using training-set-only scaling to prevent data leakage and ensure valid test set evaluation.

#### 2.3.3. Model Evaluation Strategy

For each modeling approach (XGBoost, GLM-B, GLM-Int), and for each risk tier and time window, model hyperparameters were selected using the validation set AUROC. Final performance was assessed on the held-out test set using patient-level AUROC, with 95% confidence intervals computed from 1,000 non-parametric bootstrap resamples. All note-level predictions were averaged per patient prior to evaluation to produce a single score reflecting patient-level suicide risk. Initial comparisons focus on performance across XGBoost variants: demographics-only, counts-only, and the best-performing α-weighted combination. Subsequent analyses examine performance of the penalized GLM baseline (GLM-B) and its interaction-augmented version (GLM-Int), specifically to evaluate whether inclusion of cross-modal interactions improves generalizable predictive accuracy.

Importantly, REACH-VET scores were used in two ways in this study: first, to assign patients to suicide risk tiers (low, med/moderate, high), and second, to match controls to cases within each tier. Because matching was performed on REACH-VET percentiles within tiers, REACH-VET cannot provide further discrimination between matched cases and controls. As a result, its expected AUROC within each stratum is approximately 0.5. Therefore, any model achieving AUROC > 0.5 reflects additional predictiveness beyond that captured by REACH-VET– we report the percent increase in AUROC above 0.5 as a relative improvement over this baseline.

### 2.4. Extraction and Validation of Cross-Modal Feature Interactions

#### 2.4.1. SHAP-Based Identification of Cross-Modal Interactions

For each risk tier and time window combination, we applied SHAP interaction values to the best-performing XGBoost model (as determined by α). Using the *interactiontransformer* package ^22^, we ranked pairwise feature interactions based on their global SHAP importance scores, reflecting each interaction’s average marginal contribution to model predictions across the validation set.

We focused exclusively on cross-modal interactions, defined as pairings between structured variables (e.g., demographics) and unstructured bag-of-words features. The top 100 crossmodal interactions were retained per stratum for downstream validation.

#### 2.4.2. Ranking Interactions via Negative Binomial Regression

To formally assess whether SHAP-identified interactions reflected statistically significant effect modification, we evaluated each candidate interaction using negative binomial regression models, appropriate for overdispersed count data. For each *kth* word feature *w_ij_*(*k*) in note *j* from patient *i*, and a *lth* structured feature *d_i_*^(*l*)^, we fit the following model (**Figure 1F**):

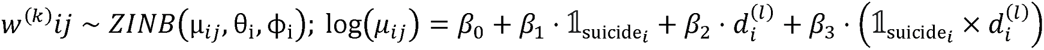

Interactions were ranked and prioritized for interpretation based on the magnitude of their effect size and the statistical significance of the interaction term (−log_10_ p-value). The coefficient *β*_3_ was statistically significant at p < 0.05. Each model included only the variables involved in the interaction term to assess evidence of effect modification. A positive interaction coefficient (*β*_3_ > 0) indicates that the relative difference in word usage between cases and controls increases with higher values of the structured variable. Conversely, a negative *β*_3_ implies that the case–control difference in word count diminishes or reverses across levels of the demographic or clinical feature. Statistical models were fit via R v4.1 (*glmmTMB*) ^25^.

### 2.4.3. Interpretation of Ranked Interactions

To characterize how word usage patterns varied as a function of suicide status and demographic context, we computed estimated marginal means (EMMs; *emmeans* R v4.1) from fitted zero-inflated negative binomial models. This allowed us to quantify, for each interaction, how the expected word frequency differed between cases and controls conditional on demographic or psychosocial strata (e.g., race, age group, housing status) ^26^. These EMMs served as the basis for interpreting the clinical or behavioral relevance of selected interactions for downstream discussion and visualization.

## 3. Results

### 3.1. Predictive Performance of XGBoost Models Across and Within Modalities

For each combination of suicide risk tier (low, medium/moderate, high) and time window (<30, <90, <270 days), we trained XGBoost models using three different feature sets: (1) structured patient characteristics only, (2) unstructured word count features only, and (3) an α-weighted mixture of both. Across all strata, models trained on structured features outperformed those trained solely on text features (**Table 4**). However, combining structured and unstructured features through an optimally tuned α led to further gains in performance in eight of nine risk-tier time window combinations, particularly in the low and medium risk tiers. This improvement was greatest for the low-risk tier when incorporating less than 270 days of EHR data for low-risk tier, which resulted in 46.2% (95% CI: 41.2–52.0) increase in predictive accuracy than REACH-VET alone. The medium-risk tier showed maximal improvement with less than 30 days of EHR data, resulting in 37.8% (95% CI: 30.6–44.8) gain in AUROC over REACH-VET alone. In both tiers, multimodal models that included demographics and word count features consistently outperformed models using demographics alone. In contrast, for the high-risk tier, combined modalities provided minimal benefit beyond structured features (**Table 4**). Structured features alone produced a 52.4% (95% CI: 40.8–62.4) beyond improvement over REACH-VET when less than 30 days of EHR data was incorporated.

### 3.2. Bridging the Performance Gap Between GLMs and XGBoost Through Validated Cross-Modal Interactions

XGBoost approaches consistently outperformed relative to baseline penalized generalized linear models (GLM-B) without interaction terms across most risk tiers and time windows, with the exception of the high-risk tier, where predictive performance was more comparable (**Table 4**). To explore whether XGBoost’s advantage stemmed from its ability to model complex dependencies, we extracted the top 100 cross-modal interactions, defined as interactions between structured (demographic) and unstructured (word count) features, using SHAP interaction values from the best-performing α-weighted XGBoost models, stratified by risk tier and time window.

**Table 4.** Test set AUROCs by risk tier and time to death (TTD) for each modeling approach: XGBoost using demographics only, word counts only, and the best-performing α-weighted combination of both; baseline penalized GLM; and GLM extended with cross-modal interactions. Includes 95% CI (2.5%-97.5%). Top models by model type (XGBoost, GLM) and risk/time are bolded to demonstrate whether performance of the integrative interaction approaches that combine structured/unstructured information– XGBoost (α*) and GLM-Int– outperform non-integrative models.

| Risk Tier | TTD (days) | XGBoost (Demo) | XGBoost (Count) | XGBoost ($\alpha^*$) | GLM-B | GLM-Int |
| --- | --- | --- | --- | --- | --- | --- |
| **High** | <30 | **0.762**<br>**(0.704–0.812)** | 0.604<br>(0.544–0.665) | 0.751<br>(0.690–0.804) | 0.707<br>(0.640–0.768) | **0.726**<br>**(0.662–0.784)** |
|  | <90 | 0.666<br>(0.603–0.724) | 0.561<br>(0.492–0.621) | **0.697**<br>**(0.630–0.757)** | 0.664<br>(0.595–0.725) | **0.677**<br>**(0.611–0.734)** |
|  | <270 | 0.681<br>(0.623–0.738) | 0.519<br>(0.451–0.578) | **0.689**<br>**(0.621–0.742)** | 0.710<br>(0.648–0.766) | **0.721**<br>**(0.661–0.775)** |
| **Moderate** | <30 | 0.667<br>(0.633–0.703) | 0.546<br>(0.511–0.581) | **0.689**<br>**(0.653–0.724)** | 0.627<br>(0.589–0.664) | **0.651**<br>**(0.614–0.686)** |
|  | <90 | 0.670<br>(0.638–0.698) | 0.616<br>(0.583–0.647) | **0.683**<br>**(0.652–0.712)** | 0.608<br>(0.579–0.637) | **0.652**<br>**(0.621–0.681)** |
|  | <270 | 0.670<br>(0.640–0.699) | 0.596<br>(0.565–0.623) | **0.694**<br>**(0.667–0.718)** | 0.594<br>(0.563–0.622) | **0.631**<br>**(0.602–0.659)** |
| **Low** | <30 | 0.644<br>(0.600–0.686) | 0.629<br>(0.587–0.673) | **0.687**<br>**(0.646–0.727)** | 0.601<br>(0.557–0.647) | **0.635**<br>**(0.592–0.681)** |
|  | <90 | 0.679<br>(0.650–0.711) | 0.611<br>(0.576–0.647) | **0.687**<br>**(0.657–0.719)** | 0.638<br>(0.605–0.674) | **0.659**<br>**(0.627–0.694)** |
|  | <270 | 0.711<br>(0.684–0.740) | 0.630<br>(0.601–0.662) | **0.731**<br>**(0.706–0.760)** | 0.648<br>(0.619–0.677) | **0.675**<br>**(0.646–0.702)** |

Among candidate cross-modal interactions, 67% showed evidence of interaction effects in negative binomial regression models (p < 0.05). Incorporating cross-modal interaction terms into the penalized GLM framework improved predictive performance relative to the baseline GLM-B models. Performance gains for resulting GLM-Int models were most pronounced in the low- and medium-risk tiers, where AUROC values approached those of the XGBoost models, closing the performance gap between XGBoost and GLM-B by 41.5% on average. In contrast, performance gains were smaller for high-risk patients from which performance was already comparable to XGBoost (**Table 4**).

### 3.3. Interpretation of Select Cross-Modal Interactions

To further elucidate the role of cross-modal interactions in suicide risk prediction, we examined a subset of statistically significant interactions using zero-inflated negative binomial regression models. These models captured how word usage varied between cases and controls depending on structured patient characteristics. **Figure 2** presents four illustrative examples drawn from different risk tiers and time windows. Additional details and interpretation are provided in the figure legend.

**Figure 2.**
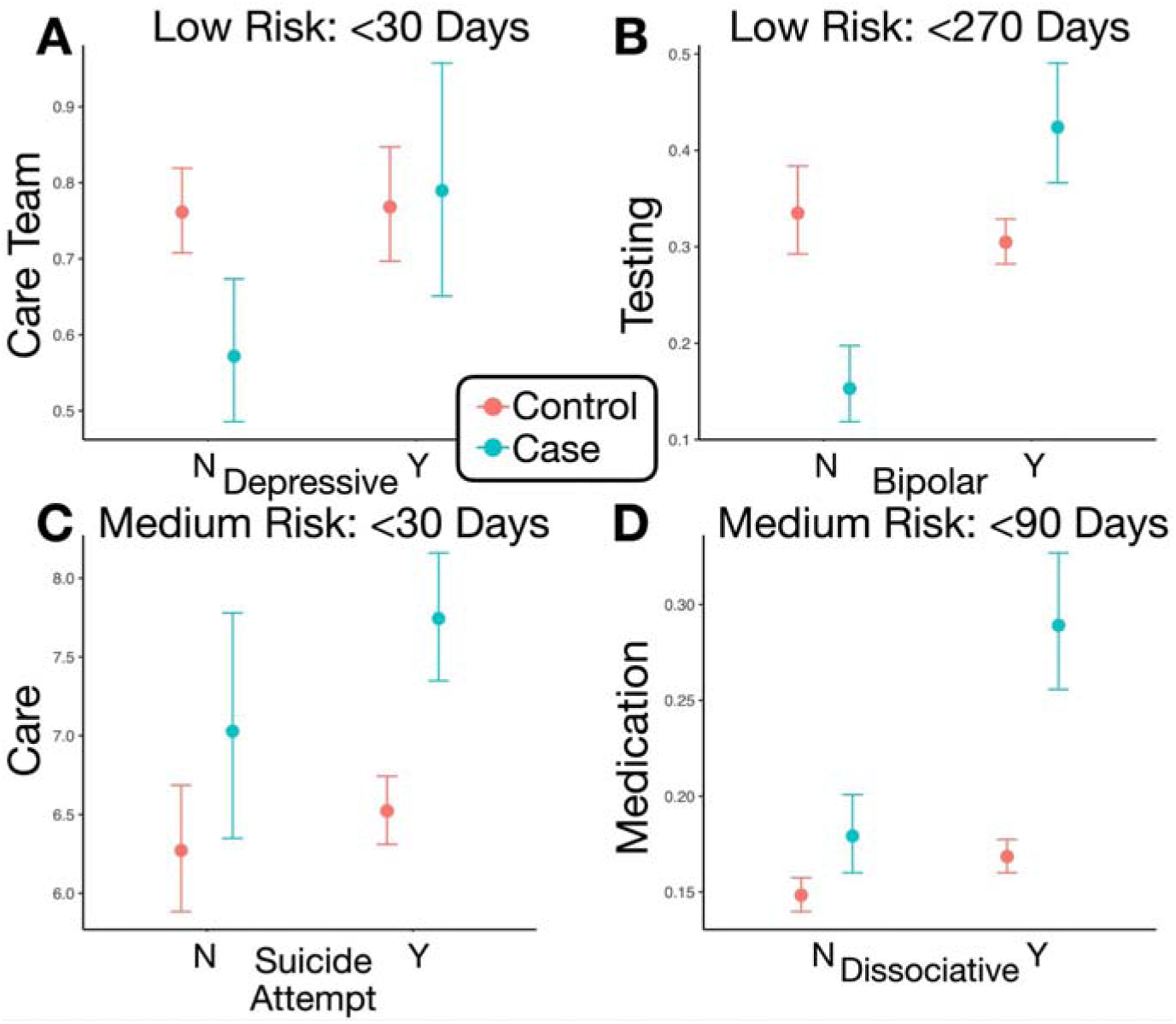
Estimated marginal means of word counts from validated cross-modal interactions, stratified by case-control status. Each panel illustrates one statistically validated interaction between a structured patient characteristic (x-axis) and an NLP-derived word feature (log word count on y-axis), estimated using a zero-inflated negative binomial regression model. Red and blue points represent controls and cases, respectively, with 95% confidence intervals. **A)** (Low Risk, <30 Days): Among patients with a depressive disorder, suicide decedents similar degree of “care team”–related language to matched controls, while “care team” was less commonly mentioned for suicide cases for non-depressive patients. **B)** (Low Risk, <270 Days): References to “testing” terms were more prevalent among cases with bipolar disorder. **C)** (Medium Risk, <30 Days): Suicide cases with a history of suicide attempt showed elevated use of “care” language in direct reference to the patient compared to controls. **D)** (Medium Risk, <90 Days): Use of medication-related terms was more common among cases with dissociative symptoms.

## 4. Discussion

Recent advancements have underscored the importance of incorporating unstructured clinical text into suicide risk prediction models ^5,27,28^. In contrast to our prior studies, which respectively investigated the incorporation of nuanced psychosocial information (SÉANCE) from clinical documents ^10,29^, derivation of risk-tier sub-populations ^8^, and identification of temporal risk fluctuation and time-sensitive risk variables ^30^, this study uniquely links these pursuits to improve personalized suicide risk efforts.

Within the current study, we found that combining structured and unstructured features using an optimally tuned α resulted in additional performance improvements, especially for patients in the low- and medium-risk tiers, leading to a 46.2% (95% CI: 41.2–52.0) and 37.8% (95% CI: 30.6–44.8) increase in predictive performance at <270 days and <30 days, respectively.

In these groups, models incorporating both demographics and word-based features consistently outperformed those using demographics alone. Conversely, in the high-risk tier, integrating unstructured data provided little to no added benefit over models relying solely on structured predictors. Notably, the integration strategy presented here (AUROCs: Low = 0.731, Medium = 0.694, High = 0.726) outperformed our prior work using SÉANCE-derived semantic variables (AUROCs: Low = 0.678, Medium = 0.658, High = 0.726).

Our findings highlight the value of integrating structured and unstructured clinical data to enhance personalized suicide risk prediction. While structured features alone provided strong baseline performance, especially for the higher-risk patients REACH-VET was designed to identify, the inclusion of unstructured word count features, when optimally weighted, yielded meaningful improvements in the low and medium risk tiers. This is noteworthy as these subgroups typically receive less care than higher-risk patients, which results in less EHR documentation and less information available for EHR-based risk algorithms to accurately assess suicide risk. This may be why the best performing model for the low-risk tier was also the model with the longest window of data inclusion that we evaluated (<270 days) whereas the medium risk tier was able to achieve maximal performance with just <30 days of EHR data. These results suggest that language patterns captured in clinical notes offer additional, personalized signals that can refine risk estimation.

The performance gap between linear models and XGBoost was largely attributed to XGBoost’s ability to capture complex cross-modal interactions. By identifying and validating these interactions, we were able to incorporate them into a penalized GLM framework, significantly narrowing the gap (∼42%) in predictive accuracy. This not only demonstrates that personalized risk and treatment factors, identified via interactions between patient demographics and clinical language within time-specific contexts, can be extracted, but that they can be successfully harnessed with interpretable modeling techniques.

Alongside our focus on cross-modal interactions and their contribution to predictive performance within the penalized modeling framework, we also developed a complementary strategy to rank interactions to aid interpretability. Given the large number of potential cross-modal interactions, examining all interactions would hinder interpretability. To avoid this concern, we focused on the following subset of statistical significant interactions with large effect sizes to illustrate clinically meaningful examples, which were also addressed in the results section.

In the first scenario, among low-risk patients who were not diagnosed with depression, suicide cases were less likely than controls to reference care team–related language in the month before death (or matching index event). This differential may suggest that these cases were less engaged in care, less supported by their providers, and less motivated by treatment, than matched controls. These domains, although difficult to measure using traditional assessment tools ^31^, have important relevance for psychotherapy outcome ^32,33^ and suicide prevention ^34^. In contrast to patients who were diagnosed with depression, and accordingly likely receive additional supportive care ^35^, the risk-burden of these non-depressed patients likely is not detected and therefore not treated ^36^. Indeed, not having a diagnosis may contribute to less follow-up care and increased care fragmentation ^36^, which are noted suicide risks ^37–39^.

In the second scenario, among low-risk patients who were not diagnosed with bipolar disorder, we found that cases had less references to testing or diagnostic evaluation than controls in the 270 days before death. Among low-risk patients who were diagnosed with bipolar disorder, we found the reverse pattern. This contrast is consistent with prior literature suggesting that bipolar patients often undergo diagnostic reassessment or increased clinical monitoring when they experience episodic changes, such as rapid cycling or mood destabilization ^40^, which can coincide with periods of heightened suicide risk. It is likely that patients who were diagnosed with bipolar disorder receive additional testing when compared to non-diagnosed patients, and that, among these patients, testing is used more frequently for patients during acute episodes ^41^, period when suicide risk is markedly high ^42^.

In the third scenario, among medium-risk patients who did and did not have prior suicide attempts, we found that cases had more references to “care” language than controls in the month before death. This may reflect intensified care coordination and clinical monitoring, interventions which are frequently triggered following a documented suicide attempt ^43,44^. It is likely that increased references to care are associated with higher symptomology and suicide risk-burdens ^41^, and this differential could help distinguish cases from controls.

Finally, in the fourth scenario, among medium-risk patients who were diagnosed with dissociative disorders, we found that cases had more references to medication than controls in the 90-days before death. Increased references to medication in this context may reflect ongoing adjustments to pharmacologic treatment, including changes in medication type, dosage, or regimen. Patients with dissociative symptoms may present complex treatment challenges and may require more frequent medication management. In some situations, repeated treatment adjustments may reflect attempts to manage escalating psychiatric symptoms in patients who ultimately progress toward suicide ^45^. These scenarios represent only a subset of the many hypothesis-generating questions that emerge when interactions are prioritized for interpretation in this way.

From a methodological standpoint, our study also reveals the feasibility of leveraging short-term language signals to inform longer-term risk prediction. Despite some temporal asynchrony between structured and unstructured predictors, these brief textual snapshots provide valuable, time-sensitive insights. In doing so, they illustrate how suicide risk fluctuates dynamically and how modeling efforts must adapt to capture this temporal specificity.

Several limitations inform directions for future work. While contextual count methods provide a scalable and interpretable proof-of-concept, they may miss deeper linguistic nuances such as negation, sarcasm, or idiomatic language. As computational resources expand within the VA’s computational infrastructure, we hope to utilize deep learning-based NLP techniques capable of modeling semantic meaning, temporal dependencies, and patient-level interactions. These models will further enable dynamic weighting of structured and unstructured features, potentially yielding temporally salient, personalized risk predictions that better inform intervention timing.

This study did not apply formal correction for multiple comparisons when evaluating interaction terms identified through SHAP ranking. Validation models included only the variables involved in each interaction and were not adjusted for additional potential confounders, as the primary objective was to screen for potential effect modification rather than estimate fully adjusted causal effects. Accordingly, identified interactions should be interpreted as hypothesis-generating and warrant confirmation in independent datasets.

We recognize the challenges and potential biases posed by data completeness in EHR data. Higher-risk patients often have more hospital visits, generate more documentation, structured information, increasing their visibility to models trained on EHR data. In contrast, low-documentation patients, often those at medium or emerging risk, remain under-identified. Our results show that language-derived features can close this gap between actual and predicted suicide risk by capturing subtleties even in sparse records, suggesting a path forward for more equitable and inclusive risk modeling for underserved populations. Further, imputation strategies may need to be adapted based on the degree of clinical documentation, though this falls outside the scope of the present study. Future work will also account for hospital site, region, provider and rural/urban locale. These results emphasize the potential for contextual, cross-modal modeling to support more tailored suicide prevention strategies.

This study provides a roadmap for shifting from population-level suicide risk stratification to individualized prediction. By uncovering how psychosocial language interacts with clinical variables across patient subgroups, we offer both practical modeling improvements and theoretical insights into the nature of suicide risk. Integrating structured and unstructured data modalities, optimizing their balance with α-tuning, and leveraging interpretable modeling techniques to capture cross-modal dynamics all support the goal of precision prevention. Particularly in lower-risk groups where signal is sparse ^8^, modeling language-derived psychosocial features alongside structured data can uncover subtle risk indicators that may otherwise go undetected. When deployed alongside existing frameworks like REACH-VET, our approach has the potential to enhance suicide detection, particularly among those who might otherwise remain invisible to standard predictive tools. Future work will expand on this foundation to identify dynamic, time-sensitive markers that further personalize risk detection and inform targeted intervention efforts.

## 5. Conclusion

This study reinforces the value of integrating unstructured clinical notes with structured electronic health record data to improve suicide risk modeling in Veterans. By appropriately balancing textual and clinical features, we identified specific patient characteristics that modified the impact of personalized language patterns on suicide risk. This approach enhanced predictive performance and establishes a meaningful baseline for future comparative studies incorporating more contextually and temporally nuanced deep learning methods.

## Author Contributions

J.J.L. and M.L. conceptualized the study, conducted formal analyses, developed methodology, performed visualization, and drafted the manuscript.

M.L. and J.G. contributed to conceptualization, funding acquisition, study oversight, and manuscript revision.

M.D. contributed to data curation, investigation, and analytic support.

L.R. contributed to data curation and data management.

S.A. provided computational resources and infrastructure support.

B.S. contributed to administrative oversight, resource acquisition, interpretation of findings, and senior leadership.

A.D., Y.Z., S.L., and W.W. contributed to data analysis, methodological support, and manuscript review.

All authors reviewed and approved the final manuscript.

## Data Availability

The data that support the findings of this study were obtained from the U.S. Department of Veterans Affairs (VA) Corporate Data Warehouse and the VA–Department of Defense Mortality Data Repository. These data contain sensitive patient information and are not publicly available. Access to VA data is subject to federal privacy regulations, institutional approvals, and data use agreements. Qualified researchers may request access through the VA Informatics and Computing Infrastructure (VINCI) or other VA data governance mechanisms. Analytic code may be made available from the corresponding author upon reasonable request.

## Ethics Declarations

This study was approved by the Institutional Review Board of the Department of Veterans Affairs at White River Junction VA Medical Center. The requirement for informed consent was waived due to the retrospective nature of the study and use of de-identified electronic health record data. All analyses were conducted in accordance with relevant guidelines and regulations governing research involving human subjects and protected health information.

